# Individual Patient Data Meta-Analysis evaluating Camostat Mesilate to Treat COVID-19 in Community Settings

**DOI:** 10.1101/2024.05.15.24307072

**Authors:** Haley Hedlin, Els Tobback, Justin Lee, Yiwen Wang, Ilaria Dragoni, Daniel C. Anthony, Kevin Dhaliwal, John Norrie, Sarah Halford, Jose Gotes, Mariana Moctezuma, Antonio Olivas-Martinez, Chaitan Khosla, Upi Singh, Jesper Damsgaard Gunst, Alonso Valdez, David Kershenobich, David Boutboul, Ole S. Søgaard, Marie-Angélique De Scheerder, Manisha Desai, Julie Parsonnet, Camostat Pooled Analysis Consortium

## Abstract

**Background:** In the COVID-19 pandemic, a number of phase II and III randomized trials were launched to evaluate the effectiveness of camostat, an orally administered TMPRSS2 inhibitor previously approved for other indications, for treating SARS-CoV-2 infections. Owing to the rapidly changing landscape during the pandemic, many of these trials were unable to reach completion. Further, methods for synthesizing data for trials that were launched and not completed were critical.

**Methods:** This study aimed to consolidate global evidence by identifying placebo-controlled, randomized trials of camostat and analyzing their collective clinical and virologic impact on SARS-CoV-2 through an individual participant data meta-analysis. We harmonized data from the included studies and utilized Bayesian statistical models to assess virologic outcomes (measured by the rate of change in viral shedding) and clinical outcomes (based on the time to the first of two consecutive symptom-free days), adjusting for age and sex.

**Findings:** The meta-analysis incorporated data from six countries, totaling 431 patients across the studies; 118 patients contributed data for the primary virologic outcome and 240 for the clinical symptom outcome. Camostat did not improve the rate of change in viral load (difference in rate of change = 0.11 Ct value/day higher, 95% credible interval 2.04 lower to 2.23 higher) or time to symptom resolution (hazard ratio = 0.87, 95% credible interval 0.51, 1.55) when compared to placebo.

**Interpretation:** In a meta-analysis prompted by a fast-changing landscape during the pandemic, we jointly synthesized evidence across multiple trials that did not meet their original recruitment goals. Despite its theoretically promising mode of action, camostat did not demonstrate a statistically significant virologic or clinical benefit in treating COVID-19, highlighting the complexity of drug repurposing in emergency health situations.

**Funding:** This work was partially supported by The Lundbeck Foundation, LifeArc, Assistance Publique Hôpitaux de Paris, anonymous donors, and awards from the National Institutes of Health.

**Research in context:** *Evidence before this study:* Camostat mesilate, a therapy widely used in Japan for over two decades to treat pancreatitis and reflux esophagitis, showed promise against SARS-CoV-2 in early laboratory and animal studies. Numerous studies evaluating camostat as a treatment for COVID-19 were launched by autumn of 2020, but later stalled due to emerging treatments that altered the equipoise for placebo-controlled trials. Among the trials that reached publication, findings were mixed.

*Added value of this study:* Our research brings a fresh perspective by comprehensively analyzing both published and previously unseen data from randomized clinical trials on camostat. By pooling data across studies, our analysis provides a more robust assessment of the effectiveness of camostat against viral and clinical outcomes than any single study could offer. Novel analytic approaches, data sharing efforts, and international collaboration during the global health emergency are additionally described.

*Implications of all the available evidence:* After thorough analysis, our study concludes that, when considering all available data, camostat does not confer a virologic or clinical advantage in the treatment of COVID-19. This conclusion underscores the importance of pooling global research efforts to build a clearer understanding of potential treatments during health emergencies.

## Introduction

Since its emergence at the end of 2019, coronavirus disease 2019 (COVID-19) has spread worldwide with clinical presentations ranging from asymptomatic to life-threatening.^1^ The rise of the severe acute respiratory syndrome coronavirus 2 (SARS-CoV-2) and the economic and social impact caused a surge of research and the rapid development of several effective vaccines as well as novel therapeutic agents. Camostat mesilate/Foipan is a serine protease inhibitor that has been licensed and extensively used in patients with pancreatitis and reflux esophagitis for more than 20 years in Japan.^2^ It is a prodrug whose metabolite 4-(4-guanidinobenzoyloxy) phenylacetate (GBPA) is the active compound, also known as FOY-251 (Midgley et al. 1994). After its original approval for the relief of abdominal pain associated with chronic pancreatitis,^2^ camostat was found *in vitro* to inhibit transmembrane protease serine type 2 (TMPRSS2), which plays a critical role in the virus life cycle of SARS-CoV-2.^3^ TMPRSS2 is expressed in the human respiratory tract and contributes to SARS-CoV-2 infection and spread by cleaving and primingthe viral spike (S) protein that subsequently binds the ACE2 receptor and allows viral entry into target cells.^4^ The potential importance of TMPRSS2 was shown by enhanced SARS-CoV-2 infection in an engineered TMPRSS2 overexpressing cell line (VeroE6/TMPRSS2) and by viral spike (S) protein priming specifically in TMPRSS2 expressing cells (Caco-2).^4,5^ Additionally, camostat was previously found to be effective in protecting mice against death due to a lethal infection by SARS-CoV, with a survival rate of approximately 60%.^6^ These studies suggested a potential mechanism of action for camostat in treating SARS-CoV-2 infection, and interest in the drug increased globally.

By the autumn of 2020, studies evaluating camostat as a potential treatment for COVID-19 had been launched.^7^ A virtual symposium on 29th October 2020 at which protocols were shared attracted over 100 participants. The landscape for treatment and prevention of SARS-CoV-2, however, rapidly changed. Remdesivir was approved for the treatment of hospitalized patients in October 2020 and monoclonal antibodies were available by the end of 2020 for outpatients; the first COVID-19 vaccine was administered on 8 December 2020. Investigators were concerned that the emerging treatments altered equipoise for placebo-controlled trials; the onset of vaccines also reduced the frequency of serious illness, the endpoint of most trials. Consequently, many of the trials investigating camostat stalled and only a few published their findings (Table 1).^8–14^ Those that have been published demonstrated inconsistent results, each focusing on different target subgroups with limited ability to draw informative conclusions.

**Table 1:**
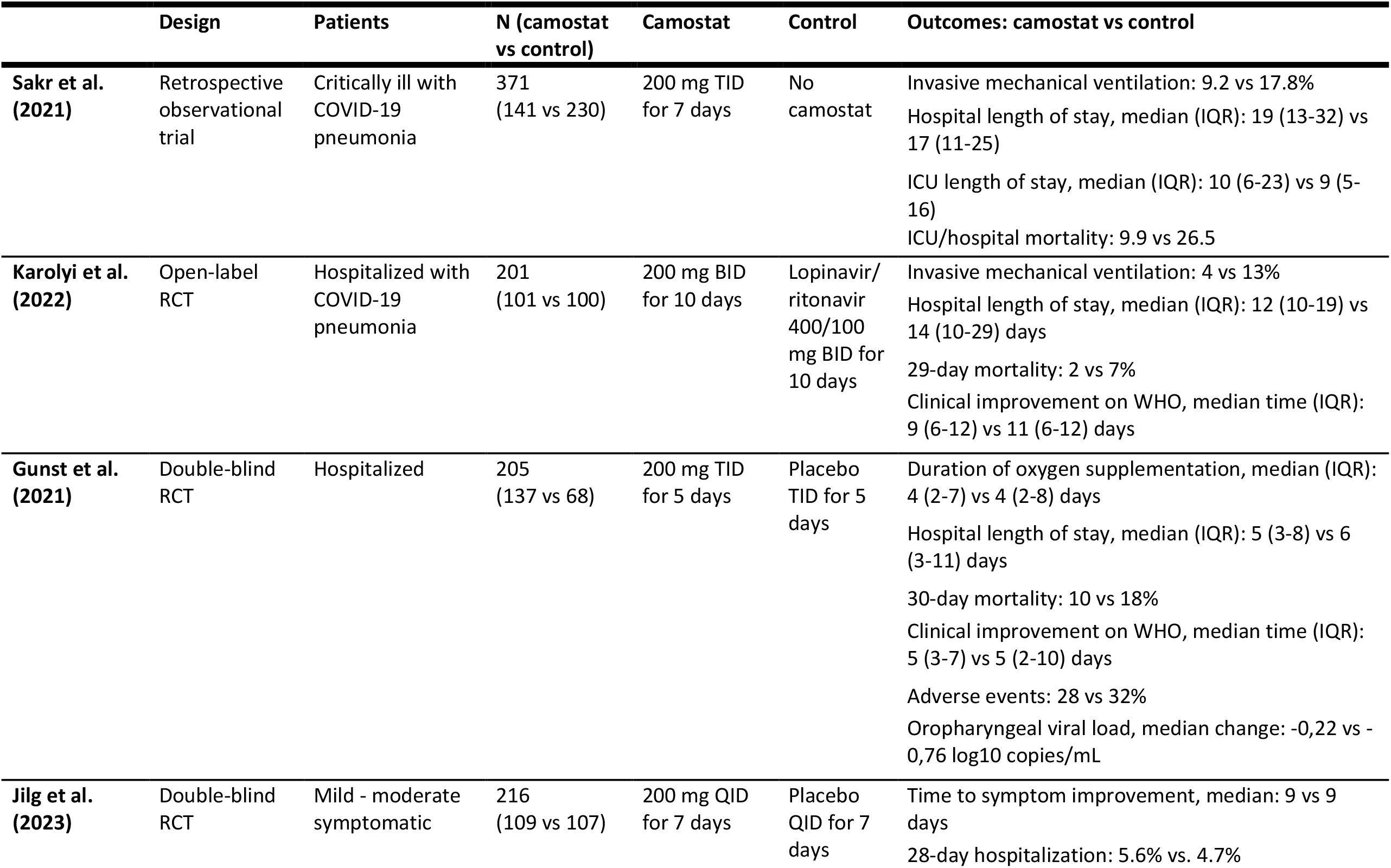
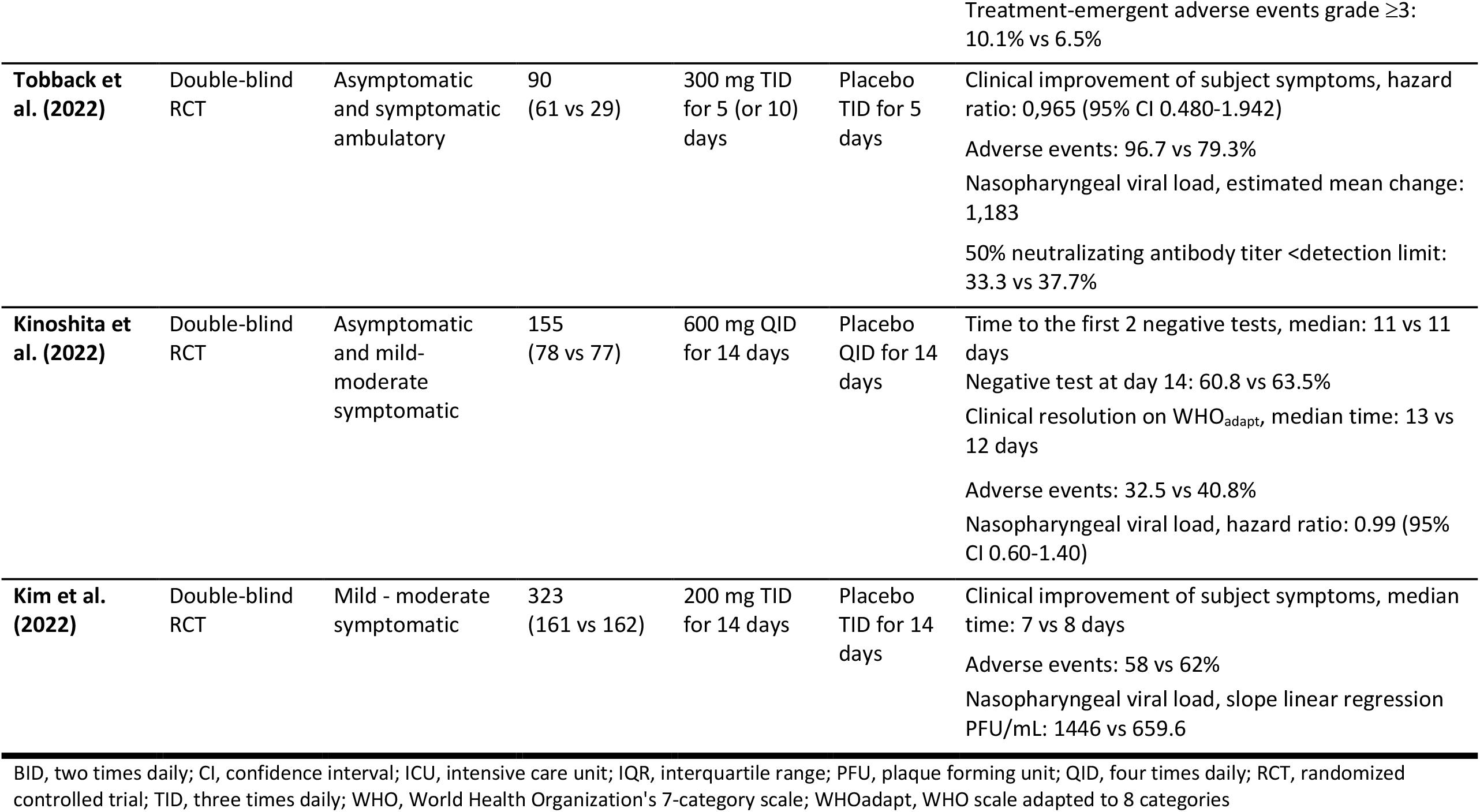
Summary of published trials evaluating camostat in COVID-19.

Because camostat remains a plausible and likely safe treatment, and because some previously useful treatments—such as monoclonal antibodies—are no longer effective, we asked investigators with ongoing, terminated, or paused trials studying camostat to participate in a individual participant data meta analysis using outpatient randomized controlled trials (RCTs). Our aim was to synthesize evidence to make a collectively stronger conclusion than could be achieved with any individual trial.^11,12,15,16^ This pooled study, which was facilitated by novel analytic methods and data sharing efforts during the global health emergency,^17–21^ assessed the efficacy of camostat in outpatients with mild COVID-19 in reducing viral shedding (viral endpoints) and symptom duration (clinical endpoints).

## Methods

Eligible outpatient studies of camostat were identified through www.clinicaltrials.gov. To be included, studies needed to be an RCT evaluating the efficacy of camostat in adult outpatients with acute SARS-CoV-2 as compared to the standard of care. The studies also needed to collect data on either viral or clinical endpoints to allow harmonization across studies in a meta-analysis. Nine studies were identified, and all were invited to participate. Out of these, 6 agreed to provide data for the meta-analysis: Stanford University (Stanford), Cancer Research UK (CRUK), Ghent University Hospital (UZ Ghent), National Institute of Health Sciences and Nutrition Salvador Zubiran (INCMNSZ), Aarhus University Hospital (Aarhus), and Assistance Publique–Hôpitaux de Paris (APHP). Table 2 describes each trial, including key inclusion/exclusion criteria, dose, and schedule of camostat administration, measurements collected, and sample size.

**Table 2:**
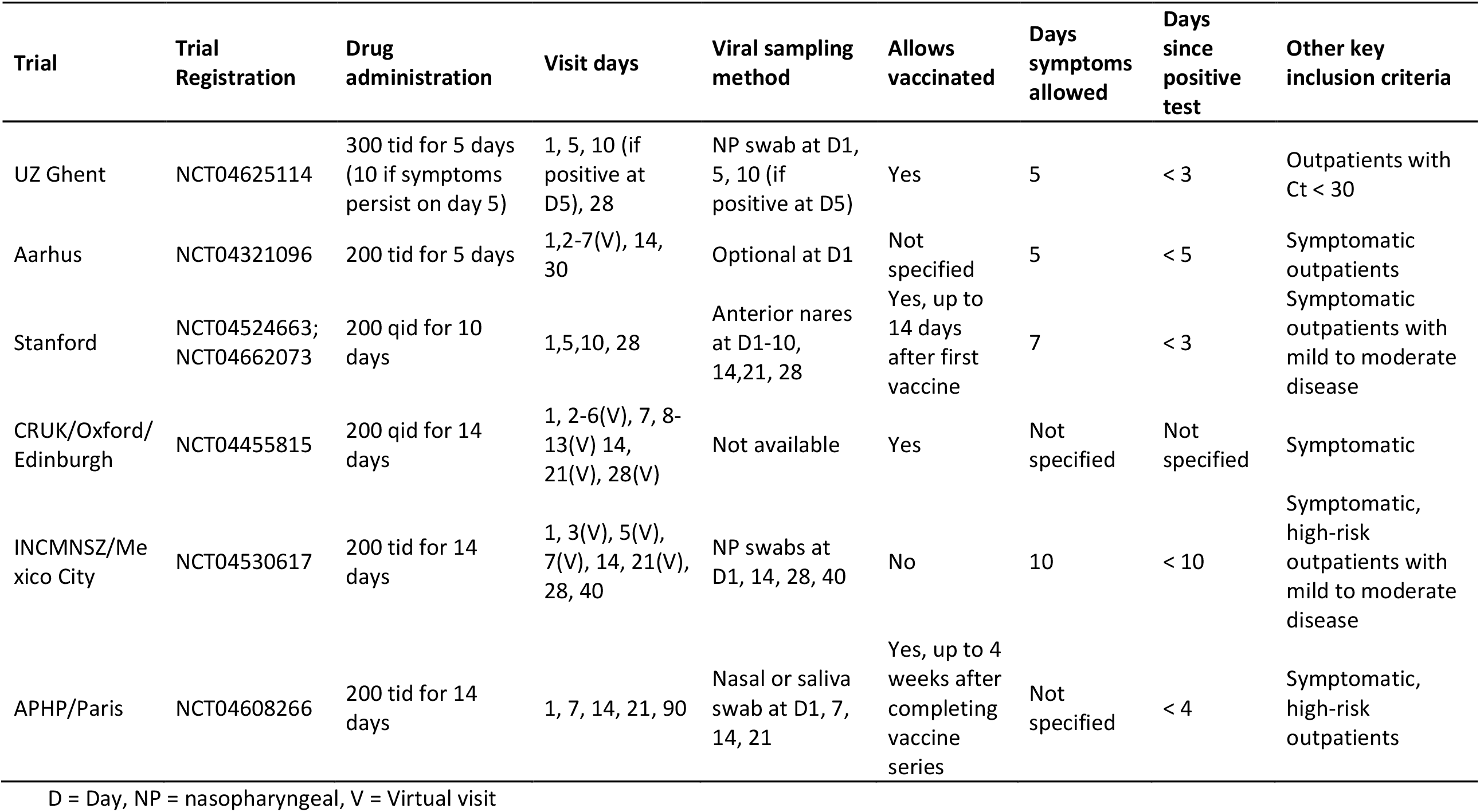
Summary of trials included in individual patient data meta-analysis.

When the investigators of the individual studies began collaborating, the first step was to compare protocols to assess feasibility of harmonizing key variables to pool individual patient data collected from the individual. After endpoint overlap across studies was assessed, we defined the objectives, endpoints, and methods appropriate to achieve our goals in a pre-specified statistical analysis plan (SAP) (see Supplemental Materials)^22^. We modeled our design after the methods of Troxel et al. in following a Bayesian monitoring approach to avoid conflicts with the type I error control in the individual study designs, as several of the included studies had paused enrollment and could possibly continue to completion.^18^

Enrolled patients had been randomized into two arms: (1) the control arm, comprised of standard supportive care in addition to placebo in all trials except CRUK where the comparator was standard supportive care alone and (2) the treatment arm, comprised of camostat in addition to standard supportive care (see Table 2 for dosage). Randomization was performed in a 1:1 ratio camostat:control in all trials except UZ Ghent, which randomized in a 2:1 ratio. Randomization at Stanford was stratified by age and sex.

### Data sharing/MoU

A Memorandum of Understanding (MoU) was put in place between all participating institutions and Ono Pharmaceutical Co., Ltd., the manufacturer and supplier of camostat. The MoU included (1) the institution serving as the data coordinating center to store, manage, and analyze the data, (2) the outline of the statistical analysis to be performed, (3) Standard Contractual Clauses to ensure compliance with the requirements of Regulation (EU) 2016/679 of the European Parliament and of the Council of 27 April 2016 on the protection of natural persons with regard to the processing of personal data and on the free movement of such data (General Data Protection Regulation, GDPR) for the transfer of personal data to a third country, (4) the UK addendum to the EU Commission Standard Contractual Clauses to provide appropriate safeguards for the purposes of transfers of personal data from the UK to a third country or an international organization in accordance with Articles 46 of the UK GDPR Law and (5) the description of the technical and organizational measures to ensure the security of data transfer and storage.

### Harmonizing Viral endpoints

One or more viral endpoints were available from four studies: Stanford, APHP, UZ Ghent and INCMNSZ. For this meta-analysis, the harmonized primary viral endpoint was rate of change in viral load as measured by cycle threshold (Ct) that is inversely proportional to viral load (the higher the value, the lower the virus, as it indicates more cycles are needed to detect virus) obtained from nasal swabs at Day 1 and 5 (UZ Ghent) and anterior nares swabs at Days 1 through 5 (Stanford). A secondary viral endpoint was an indicator for testing positive at Day 14 (anterior nares swabs from Stanford, APHP, and nasopharyngeal samples from INCMNSZ).

### Harmonizing Clinical endpoints

All studies collected clinical endpoints of some type but at varying time points. The following symptoms collected by Stanford, CRUK, UZ Ghent, and Aarhus at consistent time points were included in the symptom resolution endpoint: shortness of breath, fatigue (excluding mild fatigue), nausea, sore throat, nasal congestion, myalgia, cough (excluding mild cough), and diarrhea (not available in patients contributed by CRUK). All questionnaires used an ordinal scale to collect the severity of each symptom. The Stanford trial assessed symptoms daily through Day 28 by sending participants a link to the COVID-19 Outpatient Symptom Survey.^21^ The CRUK trial assessed symptoms via daily phone or video calls using the FluiiQ Influenza Intensity and Impact Questionnaire.^23^ A questionnaire was emailed to patients enrolled in the UZ Ghent trial through Day 14 (or to Day 28 if symptoms persisted at Day 14).^12^ Participants in Aarhus were emailed a questionnaire daily until symptoms resolved or Day 30, whichever occurred first (see Supplemental Materials for symptom surveys).

The harmonized primary clinical endpoint for the meta-analysis was time to initial symptom resolution within 14 days, defined as time from randomization until the first of two consecutive days when no symptoms reported. Patients who did not meet the symptom endpoint on their last completed survey were censored.

The secondary clinical endpoint was a composite endpoint of time to hospitalization, initiation of supplemental oxygen use, and death within 28 days from time of randomization. Patients who were never hospitalized, never received supplemental oxygen, and were alive at 28 days were censored.

### Safety

Additional secondary outcomes included adverse events (AEs) and a composite of hospitalizations, supplemental oxygen use, and death during the study.

### Statistical analyses

All analyses followed the intent-to-treat (ITT) principle and, as such, we grouped patients according to their randomized treatment and included all patients randomized by the time data were shared for analysis. All analyses were adjusted for age and sex, i.e. the randomization stratification variables used at Stanford.

All models were fit using a Bayesian framework, unless otherwise noted. The distributions and parameters in the model were refined during simulations prior to unblinding. After fitting the models, we evaluated our findings’ robustness to the choice of prior distributions, e.g. we centered the distribution of the beta coefficient at a non-zero value. For example, our primary analyses relied on uninformative priors, where the prior distribution for our parameter of interest is centered at 0. In sensitivity analyses, the center of the prior distribution for our parameter of interest would be non-zero.

To evaluate the harmonized viral endpoint, we derived the rate of change in the viral load over Days 1-5 by fitting a linear regression to each patient’s Day 1-5 data. Rate of change was compared between the two arms using a linear mixed effects model with a random intercept and random slope for trial to account for correlation between observations from the same trial and heterogeneity of effects across trials.

Odds of testing positive on Day 7 and Day 14 were evaluated by treatment arm using a generalized linear mixed-effects model with a random effect for trial, fit using frequentist methods. A separate model was fit to estimate odds ratios comparing camostat to placebo for Day 7 and for Day 14 because different studies had data available for each time point.

To evaluate the harmonized clinical endpoints, time until symptom resolution was compared between treatment arms using a two-parameter frailty proportional hazards model to account for within-trial correlation. The hazard ratio for time to clinical improvement was estimated from a Weibull model with Gamma frailties. We pre-specified that if the proportional hazards assumption is not met, we would consider a model that relaxed the proportional hazards assumption.

In secondary analyses, we used a Cox proportional hazards model, fit using frequentist methods and stratified by trial to compare the time to the first of hospitalization, supplemental oxygen use, and death. The proportional hazards assumption was visually assessed. Patients who did not experience the composite endpoint were right-censored at the end of the patient’s follow-up or Day 28, whichever was earlier. We performed the hospitalization efficacy analysis in (1) all trials collecting data on hospitalization and supplemental oxygen and (2) the subset of patients from the INCMNSZ and APHP trials.

The model parameterizations and additional details are provided in the Statistical Analysis Plan (Supplemental Materials). Estimates obtained using Bayesian methods are presented with 95% credible intervals. When fitting frequentist models, we performed two-sided tests with alpha = 0.05 and present 95% confidence intervals. All analyses were conducted in R version 4.2.1.^24^ We used the ‘rstanarm’ package to fit the Bayesian models in R.

### Role of the funding source

The funders had no role in data collection, analysis, or the decision to publish.

## Results

431 patients were included in at least one analysis. The number enrolled in individual trials ranged from 34 to 114. Viral endpoints through Day 5 were derived from 118 patients at UZ Ghent and Stanford; 75 patients from Stanford and APHP contributed viral data at Day 7; 74 patients from Stanford, INCMNSZ, and APHP had viral data available at Day 14; and 240 patients contributed symptom or other clinical data from Stanford, CRUK, UZ Ghent, INCMNSZ, and Aarhus.

The mean age of participants in the studies ranged from 39 to 56 years with a minimum age of 18 and a maximum of 84. Most studies had slightly more females with a range of 37-62% female by study (Table 3).

**Table 3:** Baseline characteristics of all patients overall and by trial.

|  | Aarhus<br>(N=71) | CRUK<br>(N=34) | APHP<br>(N=67) | INCMNSZ<br>(N=114) | Stanford<br>(N=51) | UZ Ghent<br>(N=94) | Overall<br>(N=431) |
| --- | --- | --- | --- | --- | --- | --- | --- |
| **Age** |  |  |  |  |  |  |  |
| Mean (SD) | 47.5 (14.4) | 56.1 (9.13) | 55.0 (13.0) | 46.4 (13.0) | 38.8 (13.7) | 39.3 (15.6) | 46.2 (14.9) |
| Median | 49 | 56 | 55 | 47 | 36 | 37 | 48 |
| [Min, Max] | [18.0, 84.0] | [32.0, 80.0] | [19.0, 84.0] | [18.0, 76.0] | [18.0, 66.0] | [19.0, 74.0] | [18.0, 84.0] |
| **Sex** |  |  |  |  |  |  |  |
| F | 44 (62.0%) | 19 (55.9%) | 31 (46.3%) | 71 (62.3%) | 19 (37.3%) | 52 (55.3%) | 236 (54.8%) |
| M | 26 (36.6%) | 15 (44.1%) | 36 (53.7%) | 43 (37.7%) | 32 (62.7%) | 42 (44.7%) | 194 (45.0%) |
| Missing | 1 (1.4%) | 0 (0%) | 0 (0%) | 0 (0%) | 0 (0%) | 0 (0%) | 1 (0.2%) |
| **Treatment** |  |  |  |  |  |  |  |
| Placebo | 32 (45.1%) | 18 (52.9%) | 34 (50.7%) | 56 (49.1%) | 24 (47.1%) | 30 (31.9%) | 197 (45.7%) |
| Camostat | 39 (54.9%) | 16 (47.1%) | 33 (49.3%) | 58 (50.9%) | 27 (52.9%) | 64 (68.1%) | 234 (54.3%) |

### Harmonized viral endpoint

The rate of change of Ct values through Day 5 was 0.11 higher in the control arm as compared to the camostat arm (95% credible interval (CrI): 2.04 lower, 2.23 higher), meaning that the amount of detectable virus decreased somewhat more in the placebo than in the control arm. However, this observed difference was not statistically significant (Figure 2). These findings were unchanged in a sensitivity analysis shifting the prior distribution for the parameter of interest.

**Figure 1:**
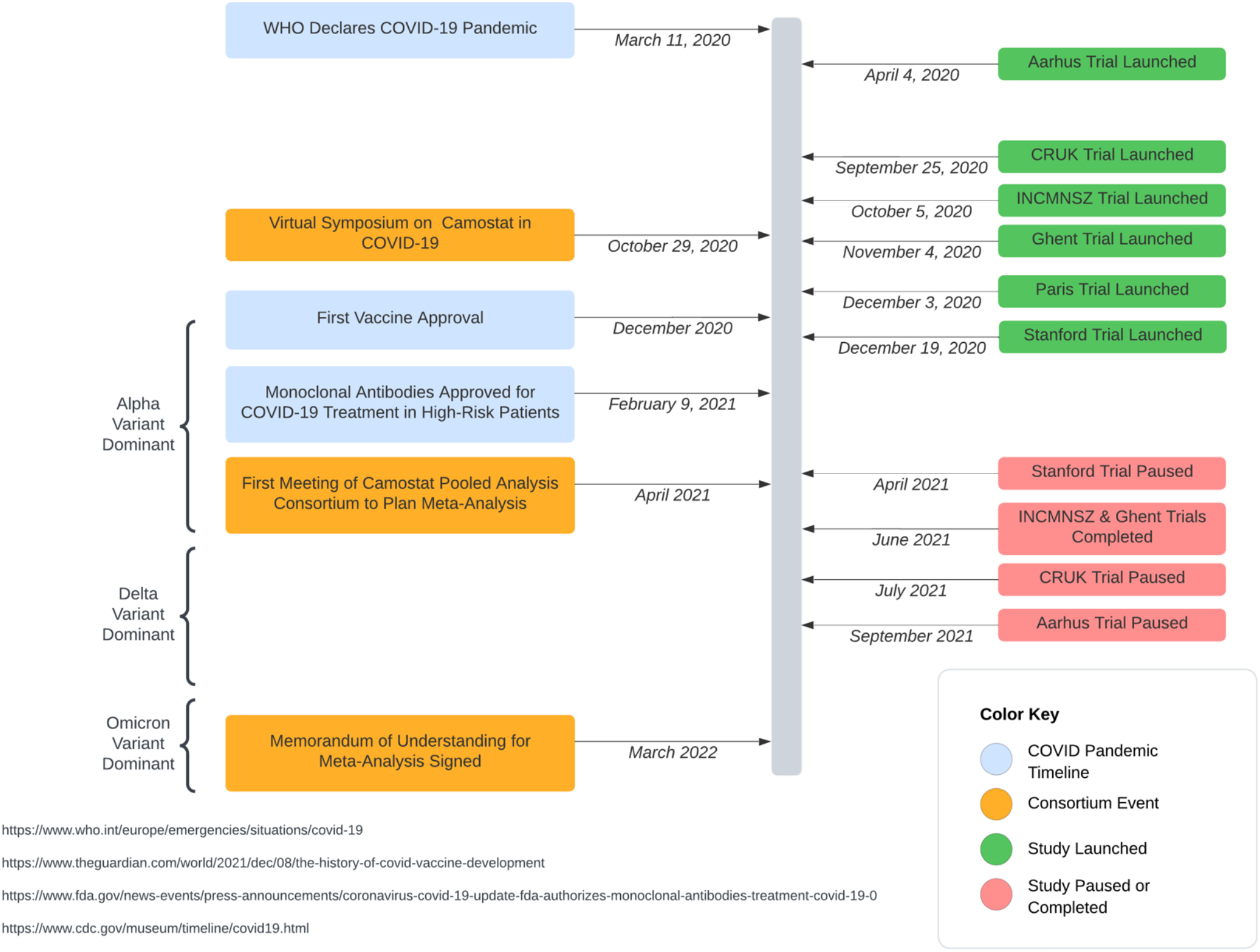
Timeline of studies in the Camostat Pooled Analysis Consortium in context of the COVID-19 pandemic

**Figure 2:**
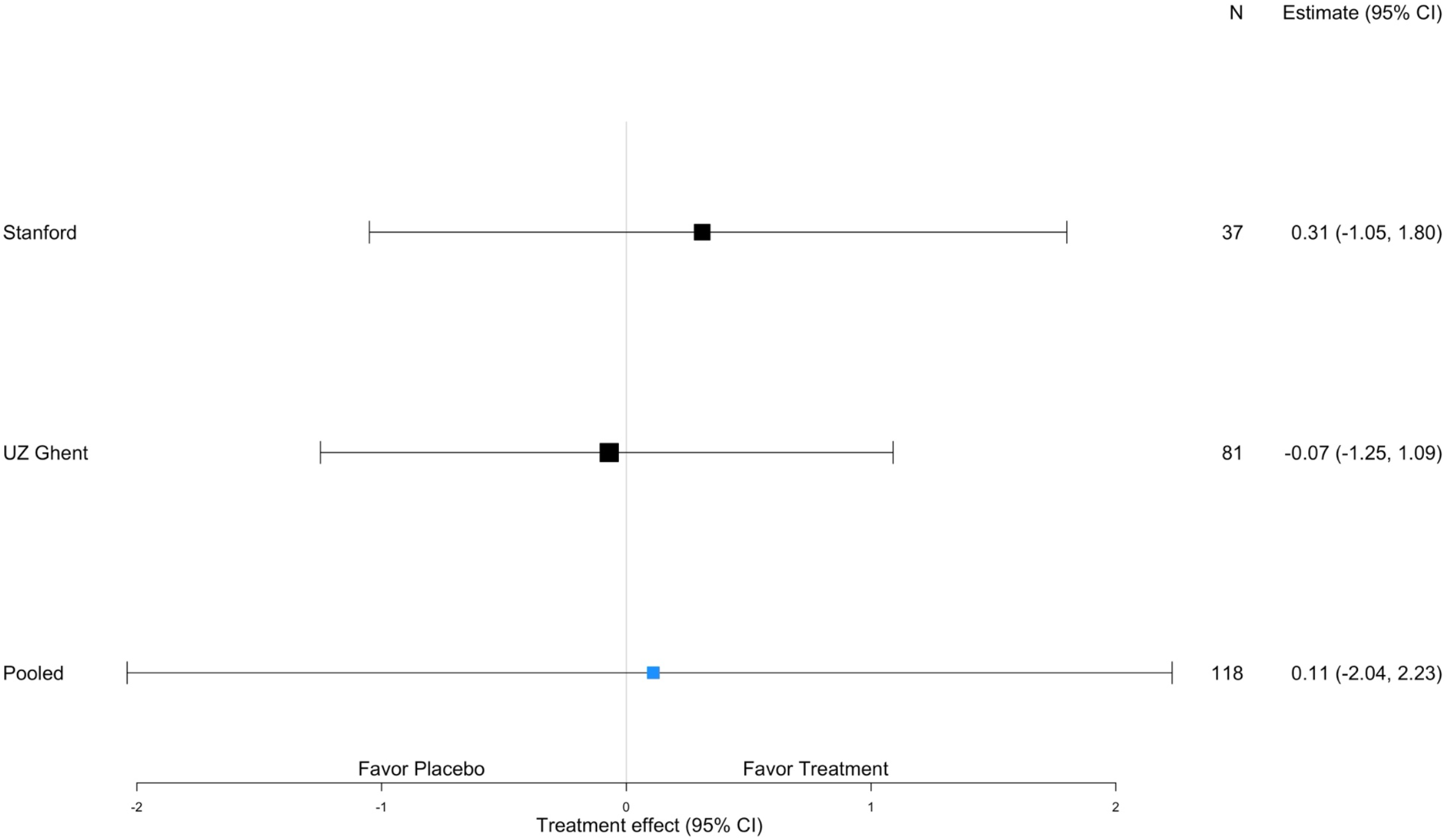
Forest plot displaying estimates and credible intervals by trial for the harmonized viral endpoint

Of 75 patients from Stanford and APHP with RT-PCR test results available at Day 7, 69 continued to amplify SARS-CoV2. The odds of a positive result were 2.20 times higher (95% CrI: 0.36 times lower, 13.4 times higher; *P*=0.40) in the camostat arm than in the placebo arm. At Day 14, 35 out of 74 patients continued to yield SARS-CoV2 RNA by RT-PCR and the estimated odds ratio of a positive test was 0.66 times lower (95% CrI: 0.25 times lower, 1.72 times higher; *P*=0.40) in the camostat arm than in the placebo arm.

### Harmonized clinical endpoints

No difference was observed in the time to symptom resolution between treatment arms (Figure 3). The estimated hazard ratio was 0.87 (95% CrI 0.51, 1.55) in favor of placebo.

**Figure 3:**
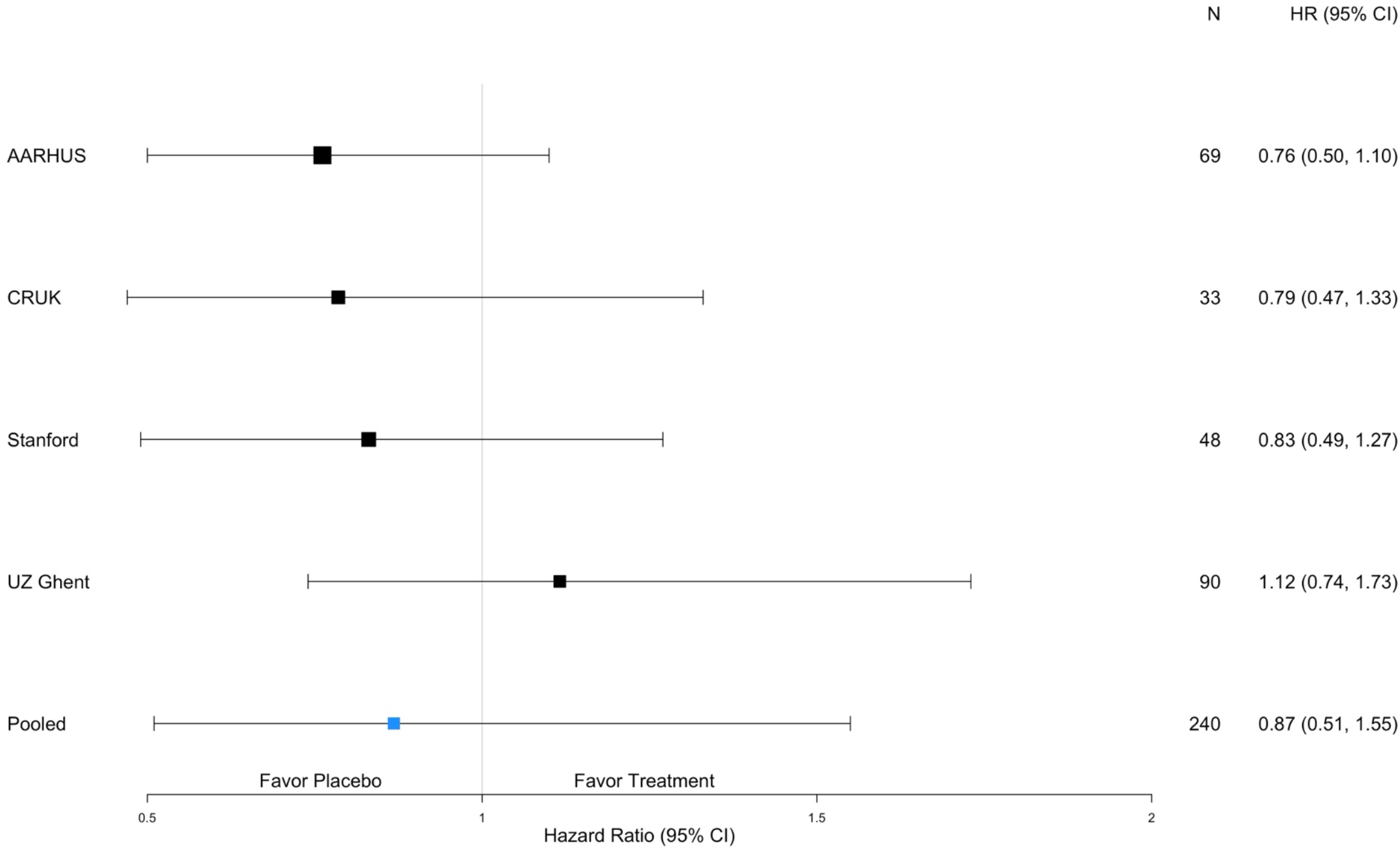
Forest plot displaying estimates and credible intervals by trial for the harmonized clinical endpoint of time to symptom resolution

### Safety

Adverse event data were available from Stanford (25 AEs), CRUK (69 AEs), UZ Ghent (257 AEs), INCMNSZ (216 AEs), and Aarhus (19 AEs). A total of 586 AEs were reported in 229 participants across the five studies. Table 4 displays the number of AEs reported in each system organ class by treatment arm.

**Table 4:**
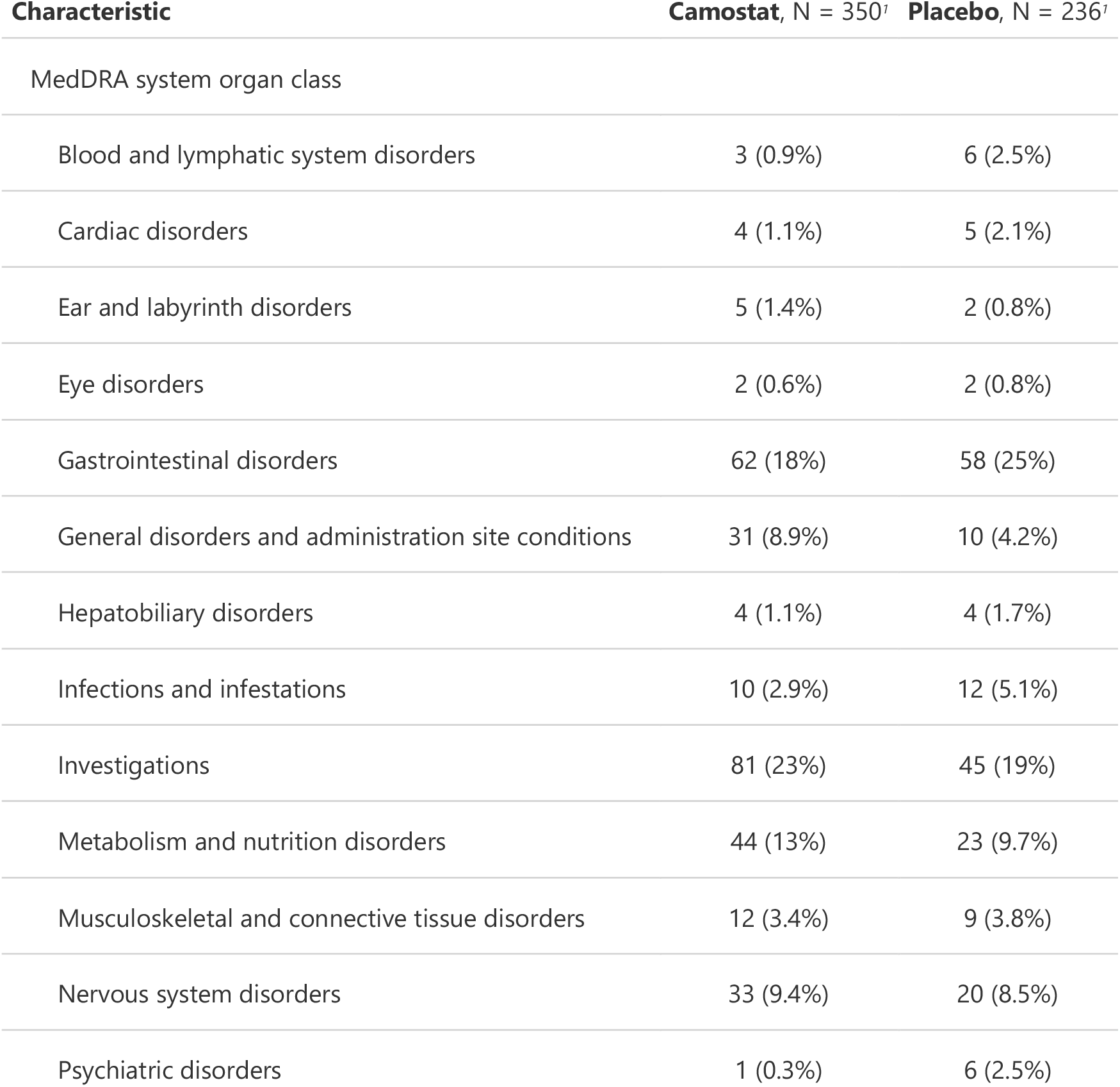
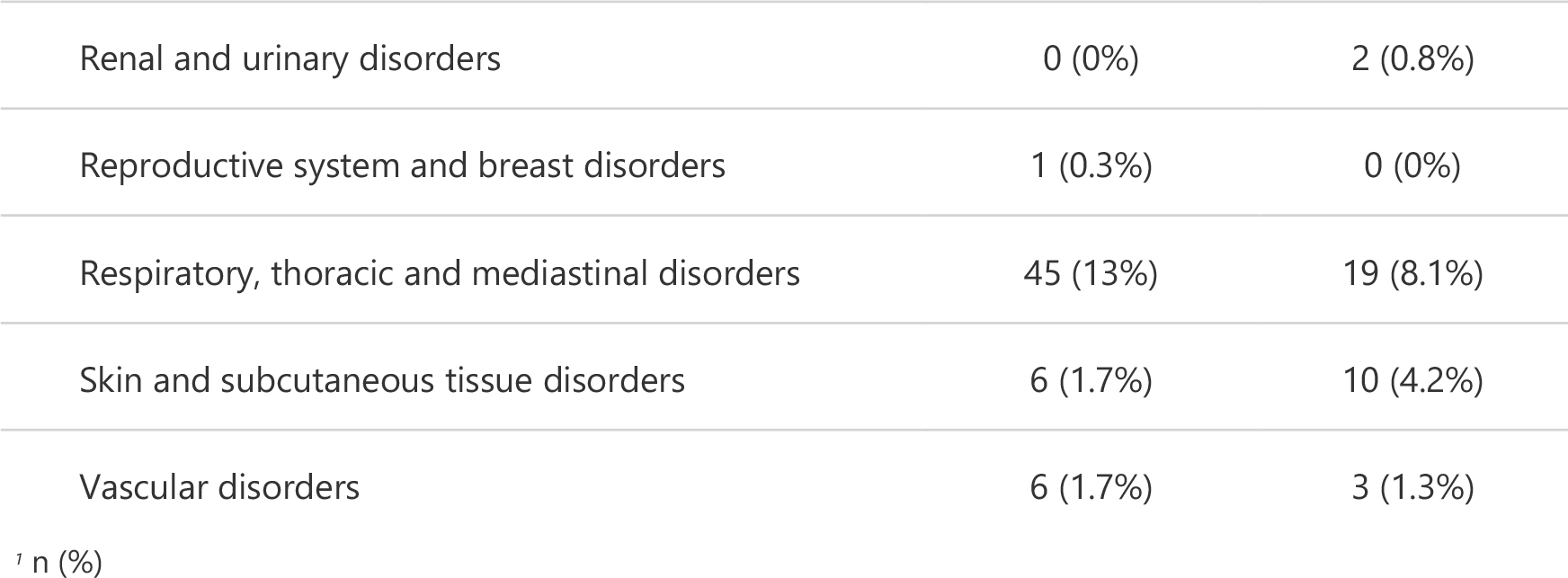
Adverse events by MedDRA system organ class and by arm.

Hospitalization was rare and no subjects died or received supplemental oxygen prior to hospitalization, so the composite endpoint is equivalent to time to hospitalization. No difference was observed in time to hospitalization between the treatment arms (HR 0.76; 95% confidence interval 0.34, 1.67). After hospitalization, there were two deaths in the camostat arm and two deaths in the control arm, all in hospitalized patients.

## Discussion

This study is the most comprehensive evaluation of the effectiveness of camostat to treat COVID-19 in community setting to date. Treatment with camostat in outpatient settings showed no difference in decline in viral shedding and no improvement in symptoms as compared to standard supportive care in patients with mild COVID-19. While there was some heterogeneity in patient populations and how the drug was administered across the trials, little heterogeneity was observed in results prior to pooling.

At the onset of the COVID-19 pandemic, numerous clinical trials were swiftly initiated, mainly targeting hospitalized patients with severe illness. However, the majority of COVID-19 patients are diagnosed and treated in outpatient settings, where there is a significant demand for treatments that can be administered conveniently to prevent worsening of the condition. The search for effective preventive and therapeutic treatments against COVID-19 in outpatient clinical trials is challenging with regard to constructional and organizational issues as well as within constantly shifting landscapes from changing virus variants to rise and fall in incidence.^25^ Recruitment slowed in all the present individual studies in parallel with the roll-out of the COVID-19 vaccination campaign along with the introduction of monoclonal antibodies and other therapeutics that perturbed equipoise and made placebo control ethically unacceptable.

This pooled analysis served as a secondary interim analysis (i.e. in addition to the interim analyses pre-specified in the individual studies) to provide guidance on how additional data can be incorporated when equipoise may be lost threatening the ability to carry out an informative trial. The paused studies specified *a priori* what their criteria would be for restarting enrollment or terminating in the SAP, a particularly important feature. At the same time the SAP was being developed, a memorandum of understanding (MOU) was being drafted between the institutions conducting the participating studies and Ono, the sponsor and supplier of camostat for some participating studies. The MOU specified that Stanford would serve as the Data Coordinating Center, the data sharing agreement to specify what data could be shared with the Stanford Data Coordinating Center, how the data would be used, and for how long the data would be stored at the Stanford Data Coordinating Center. After the SAP and MoU were finalized, each trial shared their data with the Stanford Data Coordinating Center using pre-specified data shells and data dictionaries. The effort required to gain consensus across institutions on the MoU, including the data sharing agreement adhering to the requirements of the GDPR covering the trial data collected in the European Union, was by far the most-challenging and time-consuming step of the process in this IPD meta-analysis.

The lack of efficacy of camostat on SARS-CoV-2 viral load in the present meta-analysis is in agreement with the findings in previously published RCTs where swab specimens from the nasopharynx or oropharynx were taken.^8,11–14^ Regarding the role of TMPRSS2 in viral entry into the cells, it might be thought that the inhibitory effect of camostat may especially be effective in the early phase of COVID-19. SARS-CoV-2 RNA levels peak around symptom onset and then gradually decline, supporting that administration of camostat might best be started as early as possible in the course of infection.^26^ In the present study, patients’ treatment was started within 5-7 days after symptom onset and the optimal time point for therapeutic intervention might have been exceeded. However, data from Kinoshita et al. and UZ Ghent did not show significant reductions in viral load when treatment was started at 3 days median time of symptom onset (interquartile range 0-5 and 1-4, respectively).^12,13^ In conclusion, SARS-CoV-2 viral load reduction by camostat is unlikely to be effective, even when treatment is started as early as possible in clinical practice. These results do, however, not exclude potential effects of this drug if administered as a prophylactic, for instance to close contacts of patients.

There was insufficient evidence to conclude camostat affected the time to subjective symptom resolution or the time to hospitalization in the present meta-analysis, which is in agreement with all previous studies evaluating main COVID-19 symptoms.^8,12–14^ In contrast, therapeutic benefits in terms of clinical endpoints have been described by Sakr et al. and Karolyi et al.^9,10^ These studies included moderate to severely ill patients hospitalized with COVID-19 pneumonia and concomitant therapies may also have had confounding effects on their outcomes. Taken together, camostat is unlikely to be an effective treatment for symptoms in COVID-19 outpatients.

Other trials targeting activation of S protein by TMPRSS2 with other protease inhibitors have provided insufficient evidence to conclude efficacy. Nafamostat mesilate is a synthetic protease inhibitor, which is related to camostat but has a shorter half-life and when administered intravenously in an in-patient setting, failed to reduce the length of the hospital stay or demonstrate any meaningful differences in outcomes.^27^ In a phase 3 randomized, placebo-controlled clinical trial of aprotinin, newly infected hospitalized patients with moderate COVID-19 pneumonia were given inhalation of aprotinin (2,000 KIU/day) on Day 1 of hospitalization and continued for 11 days or until discharge.^28^ Aprotinin is a pan-protease inhibitor, and inhalation of aprotinin resulted in significant shorter hospital admission compared to placebo. The trial was terminated early due to a decrease in the number of admissions, thus leaving the sample size too small to be informative. No safety concerns were reported. However, findings highlight that the inhalation route might suppress SARS-CoV-2 more effectively than systemic administration as seen with camostat or nafamostat.^29^

The present study reported substantially more adverse events in the pooled camostat arm compared to the placebo group, especially with regard to two MedDRA system organ classes, general disorders and administration site conditions and investigations. Not all studies made their safety data available, and we observed heterogeneity in reporting safety data for the reporting studies, a common problem in clinical trials.^30^

This study has numerous strengths, including its sample size, patient level data, and diversity in population as a result of participation by six trials across six different countries. The analysis highlights how a pooled analysis can be used in circumstances where equipoise may be lost and/or other extenuating circumstances can halt the progress of a clinical trial, threatening its ability to yield informative results. Unlike meta-analyses performed on published results, our meta-analysis is unlikely to be subject to the type of bias encountered with traditional meta-analyses, because we did not identify participating studies by searching published studies. Instead, we approached trials for participation by searching registries of clinical trials to see if any additional studies would be eligible for inclusion in our meta-analysis, but no additional studies were identified. Limitations of our study include the heterogeneous study designs resulting in different doses administered, inconsistent measurement schedules across trials, differing ascertainment of outcomes, and varying amounts of data available to contribute to the findings. The heterogeneity of study conduct is particularly apparent in the collection of AEs across trials. As in all meta-analyses, the interpretation of the results is limited by data from eligible trials that are unavailable at the time of the analysis.

In the future, strong consideration should be given to the idea of multiple centers joining forces to design one multi-center study of the efficacy of a given compound. This is particularly important in a pandemic, where the difficulties of initiating such studies are great, and the landscape is fast-moving and unpredictable. A multi-center study designed in collaboration at the start could very well have resulted in arriving at these findings much earlier. Furthermore, the authors here advocate for other similar efficiencies of trial design including platform or master protocols where multiple drugs can be studied simultaneously. At Stanford University, the camostat agent was indeed being studied as one of the sub-protocols in a platform protocol for this purpose.^21^ Conducting a clinical trial requires a great deal of resources to establish its infrastructure. We believe it is critical for academic and research institutions to pool resources to study multiple agents simultaneously, and that for any given agent, that it be studied in a multi-center framework. Not only will this yield huge gains in efficiency allowing us to address our questions faster and with fewer resources, but it will also allow us to achieve a higher generalizability of findings.

In summary, our team joined together in a creative manner to salvage much effort that was invested in the launching of our respective trials so that we could draw conclusions with strength. We did so in the middle of a pandemic and applied novel principles to design an overarching study that enabled our respective initial study designs to continue if needed. While harmonization of outcomes was key to the pooled analysis, we did not harmonize on triggering rules. Each trial had the flexibility to develop their own stopping rules to inform next steps. In addition, we considered both virologic and clinical endpoints. Such flexibility was critical for enabling participation by as many trials as possible. Our biggest challenge was gaining consensus in data sharing agreements that worked across our international regulatory bodies. With increased experience in collaborating on global clinical trials, we believe these challenges to inefficiencies can and should be learned and overcome.

## Data Availability

The statistical analysis plan and code are available upon request to the corresponding author (HH). The individual patient data from any specific study included in this analysis are not available through this request.

## Declaration of interests

US is an advisor to Regeneron and Gilead and her institution received research support from the National Institutes of Health, the Agency for Healthcare Research and Quality, and Pfizer, Inc.

All other authors declare no competing interests.

## Data sharing statement

The statistical analysis plan and code are available upon request to the corresponding author (HH). The individual patient data or study-level data from any specific study included in this analysis are not available through this request.

## Acknowledgments

The authors thank the participants, clinical investigators, coordinators, and contributors who have participated in the clinical trials and the individuals who helped with the data sharing and agreements required to allow us to perform this meta-analysis.

This manuscript is partially supported by the National Institutes of Health, the funding source of Stanford’s Center for Clinical and Translational Education and Research award, under the Biostatistics, Epidemiology and Research Design Shared Resource: UL1TR003142 and Stanford Cancer Institute’s Biostatistics Shared Resource (BSR): P30CA124435. The trial and work at Stanford was supported in part by anonymous donors to Stanford University and also under a Materials Transfer Agreement with Ono Pharmaceutical Co., Ltd. The trial at CRUK was funded by LifeArc. The trial in Aarhus was funded by The Lundbeck Foundation. The trial at APHP was funded by the Assistance Publique Hôpitaux de Paris. Ono Pharmaceutical Co., Ltd., provided study drug.

## Supplemental Materials

### Camostat Pooled Analysis Consortium

#### CRUK

Sarah Halford, Ilaria Dragoni, Sarah Potter, Debora Joseph-Pietras, Sawretse Leslie, Karen Hill, Susan Wan, Julie Silvester, Katie Stoddart, Helen Turner, Nigel Westwood, Neil Tremayne, Lily Elson, Isobel Hawley, Lesley Robson, Jennifer Banerjee Dhoul, Nikita Mistry, Nicola Mockler, Rachel Darby-Dowman, Stephen Nabarro, Svatopluk Svetlik, Sue Brook, Kevin Dhaliwal, Selena Harris, Salman Karim, Shruti Singh, Emma Bowen-Simpkins, Emma Ladds

#### Aarhus

Jacob Bodilsen, Lena H. Kristensen, Isik S. Johansen, Nicolai Lohse

#### Paris

David Boutboul, Lara Zafrani, Lucie Baird, Constance Guillaud, Jose Timsit, Dominique Vanjak, Georgeta Lascu, Noemie Saada, Pablo Bartolucci, Gonzalo De Luna, Raphaël Lepeule, Olivier Peyrony, Samy Ellouze, Antonella Bilgar, Ferial Belouahri, Dorothée Vallois, Anne-Claire Lehur, Nikita Dobremel, Malikhone Chansombat, Lynda Chalal, Françoise Louni, Zelie Julia, Moustafa Benafla, Christian Kassasseya, Malika Ghartouchant, Mehdi Khellaf, Chahinaz Azrar, Suella Martino, Souraya Khouider, Aurelie Sautereau, Antoine Bachelard, Pierre Leroy, Sylvain Diamantis, Lakhdar Mameri, Claire Montlahuc

#### Stanford

Julie Parsonnet, Manisha Desai, Haley Hedlin, Chaitan Khosla, Upi Singh, Hector Bonilla, Prasanna Jagannathan, Vidhya Balasubramanian, Natasha Purington, Justin Lee, Di Lu, Yiwen Wang, Savita Kamble, Catherine Ley, Dean Winslow

#### UZ Ghent

Marie-Angélique De Scheerder, Els Tobback, Liesbeth Delesie, Sophie Vanherrewege, Sophie Degroote

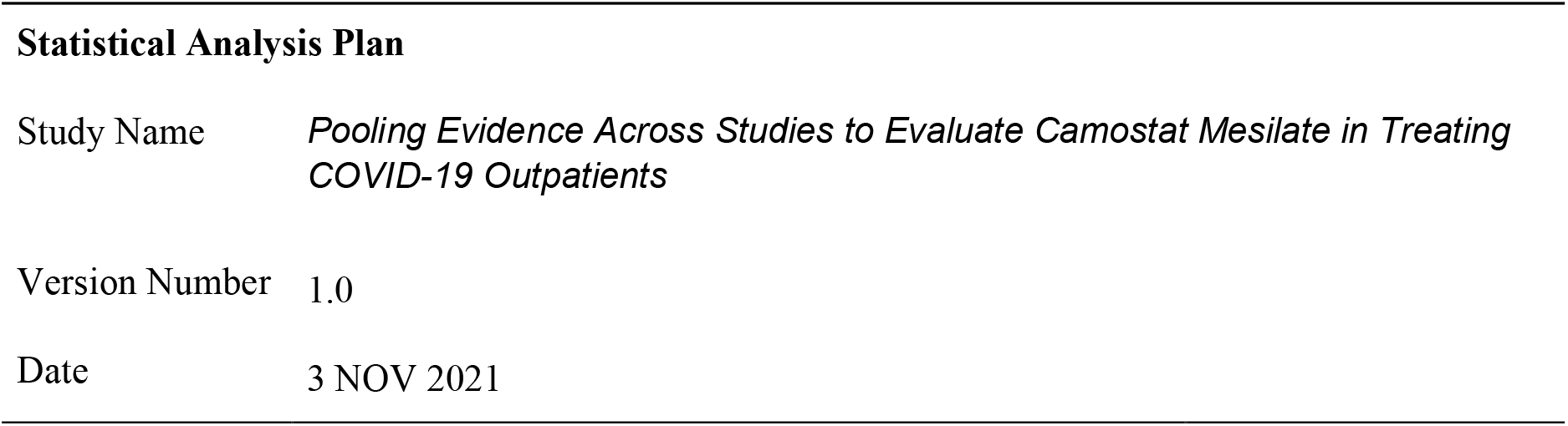

**Approved by:**

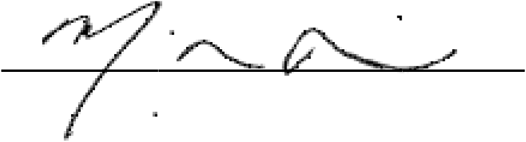

Manisha Desai, PhD

Stanford University

### Revision history

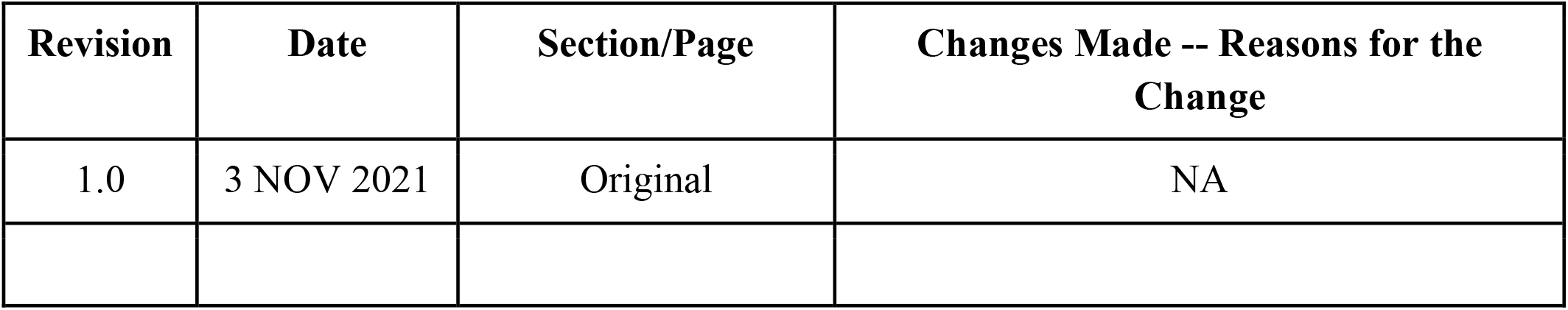

| Revision | Date | Section/Page | Changes Made -- Reasons for the Change |
| --- | --- | --- | --- |
| 1.0 | 3 NOV 2021 | Original | NA |

## Introduction

This statistical analysis plan (SAP) is a comprehensive and detailed description of the strategy, rationale, and statistical techniques that will be used in the pooled analysis.

## 1 Study details

### 1.1 Study design

This multi-institutional, individual participant data (IPD) meta-analysis intends to evaluate the efficacy of camostat mesilate in outpatients with mild COVID-19 compared with standard supportive care in reducing the viral shedding of SARS-CoV-2 virus and in treating the symptoms and clinical manifestation of COVID-19. At least 360 enrolled participants are expected to be drawn from five Phase 2 studies (Stanford University, Cancer Research UK [CRUK], Ghent Belgium University, National Institute of Health Sciences and Nutrition Salvador Zubiran [INCMNSZ], and Aarhus University Hospital) and a Phase 3 study (Assistance Publique – Hôpitaux de Paris) in this secondary interim analysis. Study participants were randomized into two arms: (1) the control arm, comprised of standard supportive care and (2) the treatment arm, comprised of camostat mesilate in addition to standard supportive care.

The primary objective of the pooled analysis is to serve as a secondary interim analysis to inform next steps as pre-specified by each individual trial team in coordination with their regulatory entities and Data and Safety Monitoring Boards. The pooled study will involve conducting two analyses to address two primary objectives: one evaluating viral shedding (where three of the four participating trials contribute data); and another evaluating clinical outcomes (where all four participating trials contribute data). The primary purpose of the virology-based objective is to examine the basic mechanisms of action on the viral shedding and viral load for camostat mesilate. The primary purpose of the symptom-based objective is to evaluate if camostat mesilate is effective in treating the symptoms and clinical manifestation of COVID-19. This is a secondary interim analysis that will inform each respective trial on next steps for the individual trial where go/no-go decisions are pre-specified among the respective study teams that are tailored to the sponsoring institution.

### 1.2 Primary viral endpoint

The primary viral endpoint is rate of change in the shedding of SARS-CoV-2 virus as measured by cycle threshold (CT) obtained from nasal swabs at Day 1 and 5 (Ghent), anterior nares swabs at Days 1 through 5 (Stanford), and saliva samples at Days 1 through 5 (CRUK).

In secondary analyses, we will compare proportion of participants who are positive at Day 7 (CRUK, Stanford, nasal swabs from Paris) and Day 14 (CRUK, Stanford, Paris, nasopharyngeal samples from INCMNSZ).

### 1.3 Primary symptom-based endpoint

The primary symptom-based endpoint is time to clinical improvement through 14 days defined as no fever for at least 48 hours AND improvement in other symptoms (e.g. cough, expectoration, myalgia, fatigue, or head ache).

## 2 Analysis plan

### 2.1 Analysis populations

All analyses will follow the intent-to-treat (ITT) principle and, as such, we will group participants according to their randomized treatment and will include all participants randomized by the data freeze, even if their data is missing.

The primary viral analysis population will include all randomized patients at Ghent, Stanford, and CRUK who were randomized and eligible to have SARS-CoV-2 viral shedding data available by the data freeze date.

The symptom analysis population will include all patients randomized prior to the data freeze date at Ghent, Stanford, CRUK, INCMNSZ, Paris, and Aarhus.

### 2.2 Descriptive analyses

Descriptive statistics (proportions for categorical variables, means, medians, standard deviations and interquartile ranges for continuous variables) will be reported for all key patient variables, including baseline and demographic characteristics and all endpoints. Data that are missing on key patient characteristics and the endpoints will be fully described, including any patterns of missingness (i.e., any relationships between missingness of a variable and patient characteristics).

A flow diagram displaying the number of patients by arm and site will be presented. Graphical tools such as histograms, boxplots, and scatterplots will be created to assess quality of data and to display patterns over time.

### 2.3 Regression covariates to account for randomization factors

For both the viral and the symptom-based endpoint, models will include participant-level factors used to generate the randomized assignments in any trial.

1. Age;
2. Sex

### 2.4 Primary viral efficacy analysis

Rate of change in the shedding of SARS-CoV-2 virus for Days 1-5 will be compared between the two arms using a linear mixed effects model with a random intercept and slope for trial to account for correlation between observations from the same trial. We will test whether Δ = 0 in the model below where *slope*_*i*_ is the derived slope for Days 1-5 for participant *i, α*_*trial*_ is a random intercept for trial, *δ*_*trial*_ is a random slope for trial, and ***X*** represents the *m* regression covariates listed in Section 2.3. The dependent variable, *slope*_*i*_, will be estimated for each participant with at least two measurements available using least squares regression.

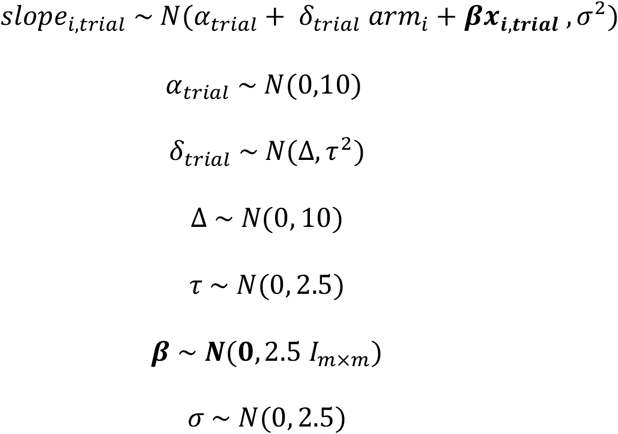

The distributions and parameters in the model will be refined during simulations prior to unblinding. For example, we may replace a prior Normal distribution with a *t* distribution or change the distribution parameters. After fitting the models, we will evaluate our findings robustness to the choice of prior distributions, e.g. we will center the distribution of Δ at a non-zero value.

### 2.5 Sensitivity analyses of viral efficacy analyses

The model described in Section 2.4 will be fit using a frequentist approach.

The model described in Section 2.4 will be fit separately within each trial using the following linear regression model. Note: ***βX*** will only be included for trials using randomization stratification. ***X*** will include the randomization variables used in each trial. The resulting estimates will be combined in a meta-analysis of the study-level results.

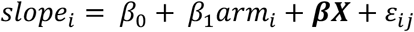

Rate of change in the shedding of SARS-CoV-2 virus for Days 1-5 will be compared between the two arms using a linear mixed effects model with a random intercept and slope for trial and participant to account for correlation between observations from the same trial or participant.

We will test whether *β*_3_ = 0 in the model below where *CT value*_*ij*_ is the CT value observed for participant *i* on day *j* for *j* = 1,2,3,4,5.

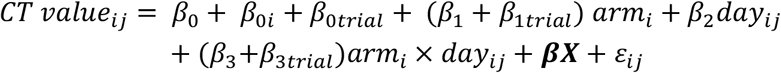

In an additional sensitivity analysis, we will fit the following model adjusting for baseline CT value where *CT value*_*ij*_ is the CT value observed for participant *i* on day *j* for *j* = 2,3,4,5 and *baseline CT value*_*ij*_ is the CT value on Day 1.

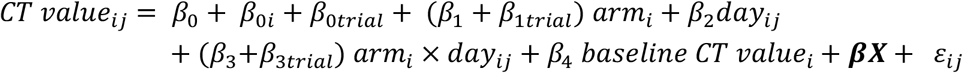

Odds of testing positive on Day 7 and Day 14 will be evaluated by treatment arm using a generalized linear mixed-effects model with a random effect for trial. *Z*_*i,trial*_ is an indicator for whether participant *i* from a trial (indicated by *trial*) tests positive.

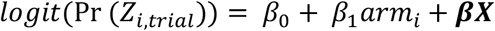

### 2.6 Primary symptom efficacy analyses

Time until clinical improvement (*y*_*i,trial*_) will be compared between the two treatment arms using a two-parameter frailty proportional hazards model to account for within-trial correlation.

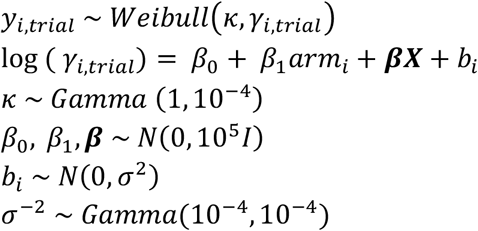

The hazard ratio for time to clinical improvement will be estimated, along with its 95% credible interval, from a Weibull model with Gamma frailties. If the proportional hazards assumption is not met, we will consider a model that relaxes the proportional hazards assumption.

The distribution of shedding cessation will be displayed using the Kaplan-Meier method, and Kaplan-Meier curves will be presented for each treatment arm overall and by trial. Time to shedding cessation at the end of the study period along with 95% confidence intervals will be presented for each treatment arm.

In a sensitivity analysis, we will consider a Cox proportional hazards model stratified by trial.

### 2.7 Hospitalization efficacy analyses

In a secondary analysis, we will consider a composite endpoint of hospitalization, supplemental oxygen use, and death through Day 28. We will use a Cox proportional hazards model stratified by trial to compare the time to the first of hospitalization, supplemental oxygen use, and death. Participants who do not experience the composite endpoint will be right-censored at the end of the participant’s follow-up or Day 28, whichever is earlier. We will perform the hospitalization efficacy analysis in 1) all trials collecting data on hospitalization and supplemental oxygen and 2) the subset of patients from the INCMNSZ and Paris trials.

### 2.8 Effect modification and subgroup analyses

A statistical interaction term between treatment arm and the baseline characteristics listed in Section 2.3 may be added to the model to test for effect modification.

Subgroup analyses may be performed according to seropositivity at baseline, defined by the presence of neutralizing antibodies against SARS-CoV-2, and asymptomatic status at baseline.

## 3 Handling of missing data

All efforts will be made to minimize instances of missing data. However, we expect some missing data will occur. Our analyses will assume data are missing at random. Participants who do not experience symptom cessation will be censored on the last day in which symptom data was reported. For other missing data, we will use multiple imputation methods that assume data are missing at random by including all baseline characteristics, treatment assignment, and reasons for missingness in the imputation model. All imputation models will be trial-specific and we will not borrow information across trials when generating imputations. This approach will be applied to any analysis involving endpoints or key variables where any missing data occurs in order to adhere to the ITT principle. Assumptions regarding missingness will be addressed in sensitivity analyses.

## 4 Interim decisions

This is a secondary interim analysis that will inform each respective trial on next steps for the individual trial where go/no-go decisions are pre-specified among the respective study teams that are tailored to the sponsoring institution.

### 4.1 Stanford University

Stanford will stop accrual and declare efficacious if posterior probability that difference between arms in Day 5 and Day 1 viral load change is greater than 3 CT is > 0.90.

Stanford will stop accrual and declare futility/harm if 1) the posterior probability that the difference between arms in Day 5 and Day 1 viral load change is greater than 1 CT is ≤ 0.50 and 2) the posterior probability that the hazard ratio for symptom resolution is 1.1 or higher is ≤ 0.50.

Stanford will continue accrual if 1) the posterior probability that the difference between arms in Day 5 and Day 1 viral load change is greater than 1 CT is > 0.50 or 2) the posterior probability that the hazard ratio for symptom resolution is 1.1 or higher is > 0.50.

### 4.2 CRUK

CRUK will continue accrual if the posterior probability that difference between arms in Day 5 and Day 1 viral load change is greater than 1 CT is > 0.90.

CRUK will stop accrual and declare efficacious if posterior probability that difference between arms in Day 5 and Day 1 viral load change is greater than 3 CT is > 0.90.

CRUK will stop accrual and declare futility/harm if posterior probability that difference between arms in Day 5 and Day 1 viral load change is greater than 1 CT is ≤ 0.90.

#### COVID-19 Outcome Symptom Scale (COSS) Daily Questionnaire

**The following questions are about how you feel and how things have been during the past 24 hours compared to your typical health. Give the one answer that comes closest to the way you have been feeling. Select “None” if you have not had this symptom**.

|  |  |  |  |  |  |
| --- | --- | --- | --- | --- | --- |
| **How do you feel compared to yesterday?** |  |  |  | ☐ I feel better than yesterday<br>☐ I feel the same as yesterday<br>☐ I feel worse than yesterday |  |
| **How do you feel compared to last week?** |  |  |  | ☐ I feel better than last week<br>☐ I feel the same as last week<br>☐ I feel worse than last week |  |
| **Cough?** |  |  |  |  |  |
| ☐ None | ☐ Mild; just a few coughs per day | ☐ Mild/Moderate | ☐ Moderate; frequent but I can tolerate it | ☐ Moderate / Severe | ☐ Severe; I am very uncomfortable |
| **Shortness of breath (difficulty breathing)?** |  |  |  |  |  |
| ☐ None | ☐ Mild; just short of breath with exercise | ☐ Mild/Moderate | ☐ Moderate; I get short of breath doing daily activities | ☐ Moderate / Severe | ☐ Severe; I feel I can't get enough air even at rest |
| **Fatigue (low energy)?** |  |  |  |  |  |
| ☐ None | ☐ Mild; I go about my day normally | ☐ Mild/Moderate | ☐ Moderate; I rest more and restrict activity | ☐ Moderate / Severe | ☐ Severe; I am staying in bed I'm so tired |
| **Headache?** |  |  |  |  |  |
| ☐ None | ☐ Mild; I can ignore it | ☐ Mild/Moderate | ☐ Moderate; I need to take medication | ☐ Moderate / Severe | ☐ Severe; it is markedly limiting my life |
| **Body Ache?** |  |  |  |  |  |
| ☐ None | ☐ Mild; I can ignore them | ☐ Mild/Moderate | ☐ Moderate; I need to restrict some activities | ☐ Moderate / Severe | ☐ Severe; they are markedly limiting my life |
| **Joint pain?** |  |  |  |  |  |
| ☐ None | ☐ Mild; I can ignore them | ☐ Mild/Moderate | ☐ Moderate; I need to restrict some activities | ☐ Moderate / Severe | ☐ Severe; they are markedly limiting my life |
| **Chest pressure?** |  |  |  |  |  |
| ☐ None | ☐ Mild; I feel it occasionally but can ignore it most of the time | ☐ Mild/Moderate | ☐ Moderate; I notice it a lot and it limits my activity | ☐ Moderate / Severe | ☐ Severe; I have bad pain and pressure that bothers me most of the time |
| **Abdominal pain?** |  |  |  |  |  |
| ☐ None | ☐ Mild; I can ignore it | ☐ Mild/Moderate | ☐ Moderate; it is limiting my activities | ☐ Moderate / Severe | ☐ Severe; it hurts a lot. I may need to see a doctor |
| **Sore Throat?** |  |  |  |  |  |
| ☐ None | ☐ Mild; I can ignore it | ☐ Mild/<br>Moderate | ☐ Moderate; it is painful to swallow and speak | ☐ Moderate / Severe | ☐ Severe; it is limiting my ability to swallow or speak |
| **Nasal Congestion?** |  |  |  |  |  |
| ☐ None | ☐ Mild; I can ignore it | ☐ Mild/<br>Moderate | ☐ Moderate; I notice it a lot | ☐ Moderate / Severe | ☐ Severe; it is markedly limiting my life |
| **Chills?** |  |  |  |  |  |
| ☐ None | ☐ Mild; I can ignore it | ☐ Mild/<br>Moderate | ☐ Moderate; I notice it a lot | ☐ Moderate / Severe | ☐ Severe; I am very uncomfortable |
| **Feeling hot or feverish?** |  |  |  |  |  |
| ☐ None | ☐ Mild; I can ignore it | ☐ Mild/<br>Moderate | ☐ Moderate; I notice it a lot | ☐ Moderate / Severe | ☐ Severe; I am very uncomfortable |
| **Runny Nose?** |  |  |  |  |  |
| ☐ None | ☐ Mild; I can ignore it | ☐ Mild/<br>Moderate | ☐ Moderate; frequent but I can tolerate it | ☐ Moderate / Severe | ☐ Severe; I am very uncomfortable |
| **Taste?** |  |  |  |  |  |
| ☐ None | ☐ Mild; tastes aren't as strong as usual | ☐ Mild/<br>Moderate | ☐ Moderate; I've noticed I can't taste certain foods | ☐ Moderate / Severe | ☐ Severe; I cannot taste my food at all |
| **Smell?** |  |  |  |  |  |
| ☐ None | ☐ Mild; smells aren't as strong as usual | ☐ Mild/<br>Moderate | ☐ Moderate; I've noticed I can't smell certain odors | ☐ Moderate / Severe | ☐ Severe; I cannot smell anything at all |
| **Diarrhea? (loose or watery stools in 24 hours)** |  |  |  |  |  |
| ☐ None | ☐ Mild; less than 3 times | ☐ Mild/<br>Moderate | ☐ Moderate; 3-6 times | ☐ Moderate / Severe | ☐ Severe; more than 6 times |
| **Nausea (feeling like you want to throw up)?** |  |  |  |  |  |
| ☐ None | ☐ Mild; I'm eating and ignoring it | ☐ Mild/<br>Moderate | ☐ Moderate; I don't want to eat and can't ignore it | ☐ Moderate / Severe | ☐ Severe; I am feeling quite uncomfortable |
| **Vomiting?** |  |  |  |  |  |
| ☐ None | ☐ Mild; only once or occasionally | ☐ Mild/<br>Moderate | ☐ Moderate; 3-4 times per day | ☐ Moderate / Severe | ☐ Severe; I am having trouble keeping food down |
| **How many times have you vomited today?** |  |  |  | _____ |  |
| **Do you have a rash?** |  |  |  | Yes / No |  |
| **The rash is (check all that apply):** |  |  |  | ☐ No rash<br>☐ Covering a small amount of my body<br>☐ Extensive covering of my body<br>☐ Involving the inside of my mouth or lips<br>☐ Itchy<br>☐ Itchy with hives |  |
| **In the past 24 hours, have you returned to your usual health (before your COVID-19 illness)?** |  |  |  | Yes / No |  |

## CR UK SPIKE1 trial

**This questionnaire will be discussed at each of the daily phone calls when your clinical research team calls you**.

- **You do not have to complete a new questionnaire each day - please try to have it with you when you are called so you can see what you will be asked and the clinical research team will record your response**.
- **Don’t worry if you don’t have it in front of you when you are called, the clinical research team will be able to read each question to you**.

**Please read each of the following questions and rate each symptom, thinking about when you felt the worst in the past 24 hours**.

1. **<u>Since this time yesterday</u>, because of COVID-19 have you had the following symptoms?**

2. **<u>Since this time yesterday</u>, has COVID-19 infection affected your ability to:**

|  | None | Mild | Moderate | Severe |
| --- | --- | --- | --- | --- |
| a. Cough | ☐ | ☐ | ☐ | ☐ |
| b. Sore throat | ☐ | ☐ | ☐ | ☐ |
| c. Headache | ☐ | ☐ | ☐ | ☐ |
| d. Nasal congestion | ☐ | ☐ | ☐ | ☐ |
| e. Feeling feverish | ☐ | ☐ | ☐ | ☐ |
| f. Body aches and pains | ☐ | ☐ | ☐ | ☐ |
| g. Fatigue (tiredness) | ☐ | ☐ | ☐ | ☐ |
| h. Neck pain | ☐ | ☐ | ☐ | ☐ |
| i. Interrupted sleep | ☐ | ☐ | ☐ | ☐ |
| j. Wheezing | ☐ | ☐ | ☐ | ☐ |
| k. Coughing up phlegm (sputum) | ☐ | ☐ | ☐ | ☐ |
| l. Shortness of breath | ☐ | ☐ | ☐ | ☐ |
| m. Loss of appetite | ☐ | ☐ | ☐ | ☐ |
| n. Loss of smell | ☐ | ☐ | ☐ | ☐ |
| o. Nausea | ☐ | ☐ | ☐ | ☐ |
| p. Loss of taste | ☐ | ☐ | ☐ | ☐ |

3. **<u>Since this time yesterday</u>, has the COVID-19 infection made you:**

**2. Since this time yesterday, has COVID-19 infection affected your ability to:**
|  | No Difficulty | Some Difficulty | Moderate Difficulty | Great Difficulty |
| --- | --- | --- | --- | --- |
| a. Get out of bed | ☐ | ☐ | ☐ | ☐ |
| b. Prepare meals / get your own food | ☐ | ☐ | ☐ | ☐ |
| c. Perform usual activities | ☐ | ☐ | ☐ | ☐ |
| d. Go out of the room you are in | ☐ | ☐ | ☐ | ☐ |
| e. Concentrate on tasks | ☐ | ☐ | ☐ | ☐ |
| f. Take care of yourself | ☐ | ☐ | ☐ | ☐ |

**3. Since this time yesterday, has the COVID-19 infection made you:**
|  | Not at all | Somewhat | Moderately | Extremely |
| --- | --- | --- | --- | --- |
| a. Irritable | ☐ | ☐ | ☐ | ☐ |
| b. Feel helpless | ☐ | ☐ | ☐ | ☐ |
| c. Worried | ☐ | ☐ | ☐ | ☐ |
| d. Frustrated | ☐ | ☐ | ☐ | ☐ |

### Symptomenvragenlijst - Questionnaire Symtoms Daily Likert

Symptomenvragenlijst - Questionnaire Symtoms Daily Likert Please indicate how much each of the following symptoms has bothered you in the last 24 hours:
|  | not at all | a little bit | somewhat | quite a bit | very much |
| --- | --- | --- | --- | --- | --- |
| 1. Runny or dripping nose | ☐ | ☐ | ☐ | ☐ | ☐ |
| 2. Congested or stuffy nose | ☐ | ☐ | ☐ | ☐ | ☐ |
| 3. Scratchy or itchy throat | ☐ | ☐ | ☐ | ☐ | ☐ |
| 4. Sore or painful throat | ☐ | ☐ | ☐ | ☐ | ☐ |
| 5. Swollen throat | ☐ | ☐ | ☐ | ☐ | ☐ |
| 6. Difficulty swallowing | ☐ | ☐ | ☐ | ☐ | ☐ |
| 7. Teary or watery eyes | ☐ | ☐ | ☐ | ☐ | ☐ |
| 8. Sore or painful eyes | ☐ | ☐ | ☐ | ☐ | ☐ |
| 9. Eyes sensitive to light | ☐ | ☐ | ☐ | ☐ | ☐ |
| 10. Trouble breathing | ☐ | ☐ | ☐ | ☐ | ☐ |
| 11. Chest congestion | ☐ | ☐ | ☐ | ☐ | ☐ |
| 12. Chest tightness | ☐ | ☐ | ☐ | ☐ | ☐ |
| 13. Dry or hacking cough | ☐ | ☐ | ☐ | ☐ | ☐ |
| 14. Wet or loose cough | ☐ | ☐ | ☐ | ☐ | ☐ |
| 15. Headache | ☐ | ☐ | ☐ | ☐ | ☐ |
| 16. Head congestion | ☐ | ☐ | ☐ | ☐ | ☐ |
| 17. Sinus pressure | ☐ | ☐ | ☐ | ☐ | ☐ |
| 18. Felt dizzy | ☐ | ☐ | ☐ | ☐ | ☐ |
| 19. Felt lightheaded | ☐ | ☐ | ☐ | ☐ | ☐ |
| 20. Lack of appetite | ☐ | ☐ | ☐ | ☐ | ☐ |
| 21. Felt nauseated | ☐ | ☐ | ☐ | ☐ | ☐ |
| 22. Stomach ache | ☐ | ☐ | ☐ | ☐ | ☐ |
| 23. Sleeping more than usual | ☐ | ☐ | ☐ | ☐ | ☐ |
| 24. Difficulty staying asleep | ☐ | ☐ | ☐ | ☐ | ☐ |
| 25. Difficulty falling asleep | ☐ | ☐ | ☐ | ☐ | ☐ |
| 26. Body aches or pains | ☐ | ☐ | ☐ | ☐ | ☐ |
| 27. Weak or tired | ☐ | ☐ | ☐ | ☐ | ☐ |
| 28. Chills or shivering | ☐ | ☐ | ☐ | ☐ | ☐ |
| 29. Felt cold | ☐ | ☐ | ☐ | ☐ | ☐ |
| 30. Felt hot | ☐ | ☐ | ☐ | ☐ | ☐ |
| 31. Sweating | ☐ | ☐ | ☐ | ☐ | ☐ |
| 32. Felt uncomfortable | ☐ | ☐ | ☐ | ☐ | ☐ |
| 33. Abnormal sense of smell or taste | ☐ | ☐ | ☐ | ☐ | ☐ |

Please indicate how many times you had each of the following symptoms in the last 24 hours:
|  | zero times | 1 time | 2 times | 3 times | 4 times or more |
| --- | --- | --- | --- | --- | --- |
| 34. Vomiting | ☐ | ☐ | ☐ | ☐ | ☐ |
| 35. Diarrhea | ☐ | ☐ | ☐ | ☐ | ☐ |
| 36. Sneezing | ☐ | ☐ | ☐ | ☐ | ☐ |
| 37. Coughing | ☐ | ☐ | ☐ | ☐ | ☐ |
| 38. Coughed up mucus or phlegm | ☐ | ☐ | ☐ | ☐ | ☐ |

39. Highest temperature in the last 24 hours: ____°C (1 decimal place)

## Notes

### Funding Statement

This work was partially supported by The Lundbeck Foundation, LifeArc, anonymous donors, and awards from the National Institutes of Health. The funders had no role in data collection, analysis, or the decision to publish.

### Author Declarations

The ethics committees/IRBs of all individual studies gave ethical approval for this work.

